# Alpha-gal Syndrome Symptom Profiles and Diagnostic Experiences Among Farmer and Ranchers

**DOI:** 10.64898/2026.04.14.26349898

**Authors:** Alexandrea M. Welch, Cheryl L. Beseler, Shaun T. Cross

**Affiliations:** Department of Environmental, Agricultural, and Occupational Health, College of Public Health, University of Nebraska Medical Center, Omaha, NE, USA; Central States Center for Agricultural Safety and Health, University of Nebraska Medical Center, Omaha, NE, USA

**Keywords:** Alpha-gal syndrome, farmers and ranchers, agricultural workers, tick-borne diseases

## Abstract

**Purpose:** Alpha-gal syndrome (AGS) is an emerging health issue. This syndrome, caused by the bites of ticks, induces allergic reactions to the sugar molecule galactose-α-1,3-galactose after exposure to non-primate mammalian meat and other byproducts. Agricultural workers spend significant time outdoors placing them at an increased risk for tick bites and tick-borne diseases, like AGS. This study aimed to characterize farmers and ranchers’ prior knowledge, symptomology, and diagnostic experiences with AGS.

**Methods:** We conducted a cross-sectional survey of more than 200 farmers and ranchers with a self-reported AGS diagnosis. The survey captured farmers and ranchers’ experiences related to prior knowledge and experience with tick bites and AGS, reported symptoms, and obtaining a diagnosis.

**Findings:** A total of 201 respondents across 26 states participated in the survey, with the majority from Missouri and Oklahoma. We identified four distinct symptom clusters, with the most reported symptoms being abdominal cramping, diarrhea, itchy skin, and nausea. Women more often reported gastrointestinal discomfort, and men were more likely to be in the mild symptom category. On average, participants reported 2.98 medical provider visits before receiving a diagnosis, most being diagnosed by general practitioners and allergists.

**Conclusions:** No previous studies have focused on the symptom and diagnostic experiences of farmers and ranchers with AGS. Capturing such data is essential as these workers may experience unique occupational challenges following AGS diagnosis. The diagnostic experience data support a continuing need to educate and empower AGS patients and providers, especially agricultural workers and providers serving rural communities.

## INTRODUCTION

Ticks are rapidly expanding into new geographic areas, in part due to climate change. With the expansion of these ticks, the burden of tick-borne diseases is steadily growing every year.^1–3^ Among these diseases is an allergy known as alpha-gal syndrome (AGS) with more than 34,000 confirmed cases; however, the estimated burden could be as high as 450,000 people in the United States.^4^ AGS is a delayed allergic reaction (three to six hours) to the sugar molecule galactose-α-1,3-galactose (alpha-gal) after exposure to non-primate mammalian byproducts, such as meat (including cattle, pig, sheep), dairy products, and certain medications.^5^ Current diagnosis for AGS can be complex as it 1) has atypical presentation of allergic reactions and 2) requires both a history of allergic reactions to mammalian by-products and a detectable threshold of immunoglobulin E (IgE) antibodies.^6^

Development of AGS is associated with tick bites, and within the United States, the most common tick bites associated with the syndrome are those by the lone star tick (*Amblyomma americanum*), though not exclusively.^7–10^ Not only is the geographic range of the lone star tick rapidly expanding, but these ticks are documented to be aggressive human biters across all life stages (larvae, nymph, adult).^11,12^ Accounting for both geographic expansion and their aggressive host-seeking behavior, there is increased likelihood of individuals to acquire lone star tick bites. Biting frequency is important as multiple tick bites have been linked to increased risk of AGS development.^13^

Though increased expansion of the lone star ticks generally increases risk of individuals for bites and potentially AGS, the risk burden is not equally shared among all. For example, as individuals spend more hours per week outside they are at an increased risk for tick exposure.^14^ Due to the nature of their work, outdoor occupational workers spend significant time outdoors placing them at an increased risk for tick-borne diseases, including within agricultural workers.^15–18^ In context of AGS, a case-control study highlighted that AGS diagnosed individuals were more likely to report to have 5 or more acres around their home, have shrubs or brush, have farmland, and have woods around the home; these serve as an environmental proxy for tick bite exposures.^19^ Given these heightened tick exposures risks due to their occupation and environment, agricultural workers may also be at a heightened risk for AGS.

Outside of increased environmental exposure to tick bites and heightened AGS risks, farmers and ranchers may also encounter barriers in obtaining a diagnosis. Individuals in rural communities, such as farmers and ranchers, are disproportionately affected by limited access to medical care.^20,21^ This limited healthcare may exacerbate current challenges in AGS diagnosis as medical provider knowledge and awareness remains low. A recent survey of 1,500 medical providers reported 42% had never heard of AGS.^22^

With these increased exposure risks and potential diagnostic challenges in agricultural occupations, there is a critical need to understand how AGS affects worker health and safety. There is increased regularity of journalism articles on the burdens of AGS among farmers and ranchers, but to date no peer-reviewed study exists that explicitly captures the symptom and diagnostic experiences. To address this gap in knowledge, this study aims to characterize farmer and rancher’s experience with AGS, by enlisting a cohort of >200 farmers and ranchers diagnosed with AGS in a cross-sectional survey. This study aimed to characterize these farmers and ranchers’ experiences related to their prior knowledge and experience with tick bites and AGS, their reported symptoms, and the process of obtaining a diagnosis.

## METHODS

### Sample

Participants were predominantly recruited from an AGS Facebook Group focused on Farmers and Ranchers with AGS. The recruitment post was shared multiple times through individual personal Facebook pages; thus we cannot establish a definitive response attributed to this group alone. Additional dissemination and recruitment occurred using a listserv network of farmers and ranchers available through the Central States Center for Agricultural Safety and Health. Individuals who had been diagnosed with AGS were invited to complete a brief questionnaire asking about their experiences with AGS. Survey inclusion criteria required participants to have an AGS diagnosis from a medical provider and/or diagnostic lab. Participants who did not report having an official diagnosis from a medical provider and/or diagnostic lab were not eligible to complete the survey. A total of 208 participants completed the survey, but respondents who did not report a primary occupation in farming or ranching were removed from the final analysis. Following removal of responses by criteria, our final sample size was 201 participants. The survey questions can be found in **Appendix 1**. The study was deemed exempt by the University of Nebraska Institutional Review Board (0562-25-EX).

### Measures

Basic demographics included sex (female = 0, male = 1), age (continuous variable), and type of agricultural work engaged in (farm or ranch). The survey asked a series of yes/no questions about whether respondents regularly check for tick bites, remembered being bitten by a tick, experienced multiple bites, were familiar with AGS, knew that tick bites could cause a food allergy, and whether they thought their symptoms were food-related. We further asked about the presence or absence of 12 symptoms known to be a result of being infected. The twelve symptoms asked about specifically came from previously published medical reports and included abdominal cramps, congestion, diarrhea, difficulty breathing, difficulty swallowing, tachycardia, hives, itchy skin, nausea, rash, swelling, and vomiting. Four additional symptoms were added due to being reported in the open-ended “other” category in the survey. These respondent-derived symptoms were fatigue, neurological symptoms (dizziness, headaches, fainting, brain fog, seizures), pain in muscles and joints, and anaphylaxis. We asked about the frequency of experiencing symptoms in four categories (daily, weekly, monthly, yearly). Lastly, we asked about the experience of obtaining a diagnosis of AGS including how many medical appointments were required to obtain a diagnosis (count variable) and what type of doctor made the diagnosis (open-ended question).

### Statistical analysis

*Descriptive statistics*. Numbers and frequencies were calculated for the demographic and binary responses (0=no, 1=yes) related to perceptions and knowledge of AGS, and the types of symptoms experienced. To explore whether certain symptoms occurred together, a multilevel latent class analysis (MLCA) was used to test for clustering of symptoms within individuals. Symptoms within individuals ranged from only one symptom to up to thirteen symptoms. With such a wide spectrum of symptoms, we hypothesized that certain types such as allergic reactions might separate out from gastrointestinal issues. We also considered the possibility that some individuals were characterized by few symptoms and some by a broader range of symptoms as a measure of severity.

*Multilevel latent class analysis (MLCA)*. To understand the grouping of symptoms, the MLCA tested one to six classes using the MLF maximum likelihood estimator and a logit link. Akaike Information Criterion (AIC), Bayesian Information Criterion (BIC), Bootstrap Likelihood Ratio Test (BLRT), and entropy were used to assess model fit. Low values of AIC and BIC suggest a better model fit than higher values. The BLRT tests whether k classes are a better fit than k-1 classes and a p-value <0.05 indicates k classes provides a better fit to the data. Entropy measures the quality of the classification and values greater than 0.80 indicate a good fit to the data.^23^ We used 500 random starting values at the initial stage and 50 at the final optimization stage to assure that the maximum likelihood estimate could be replicated and that we were estimating a global maxima. The MLCA analyses was conducted in MPlus Version 8.5.

*Correlates of symptom classes*. We used the chi-square test to assess for differences in the four classes by sex, awareness of AGS, suspected to be food-related, and having a strong reaction to tick bites. Variables that showed a significant association to class were used in a generalized logits model treating the class outcome as a nominal variable with the allergy class as a reference group. Further, chi-square tests were used to assess which symptoms were statistically associated with the classes of individuals in the MLCA.

*Assessment of the diagnosis experience*. Lastly, we used a Poisson model, adjusted for overdispersion, to identify the classes of symptoms that might be associated with having a higher number of medical provider visits in order to obtain a diagnosis. We hypothesized that the more nonspecific the symptoms, the greater the number of medical visits required to obtain an order for an AGS test. These models were adjusted by confounding variables that were associated at p<0.05 with the class of the symptoms and the number of medical provider visits. We examined the types of providers that made the diagnosis by grouping the responses provided by respondents into general medical doctors, medical specialists, and other types of medical providers (non-physician). Aside from the MLCA, all analyses were conducted in SAS 9.4 (SAS Institute, Cary, IN). Graphs were produced using ggplot2 in R version 4.3.3. Results were considered significant at alpha < 0.05.

## RESULTS

### Characteristics of the AGS sample and their experiences with tick bites

The 201 respondents were 74.1% female and primarily worked on a farming operation (61.7%) (**Table 1**). The overall age of the sample of 201 participants was 49.7 (SD=13.7). Nearly all of the respondents regularly checked for tick bites (95.0%), 77% recalled having been bitten, and 87% reported having had multiple bites. Nearly half (46.8%) knew about AGS prior to their diagnosis and 68% had heard that tick bites could cause a food allergy. Half of respondents suspected that their symptoms were food-related. Greater than 85% experienced symptoms daily or weekly, although 15% reported monthly or yearly symptoms. Overall, the respondents were diligent about checking for tick bites and educated about the adverse effects of tick bites. Responses came across the continental United States (**Figure 1**), but the distribution appeared to generally align with the distribution of the current range of the lone star tick.^24^ Most respondents were from Missouri and Oklahoma with 47 and 26 responses, respectively.

**Figure 1.**
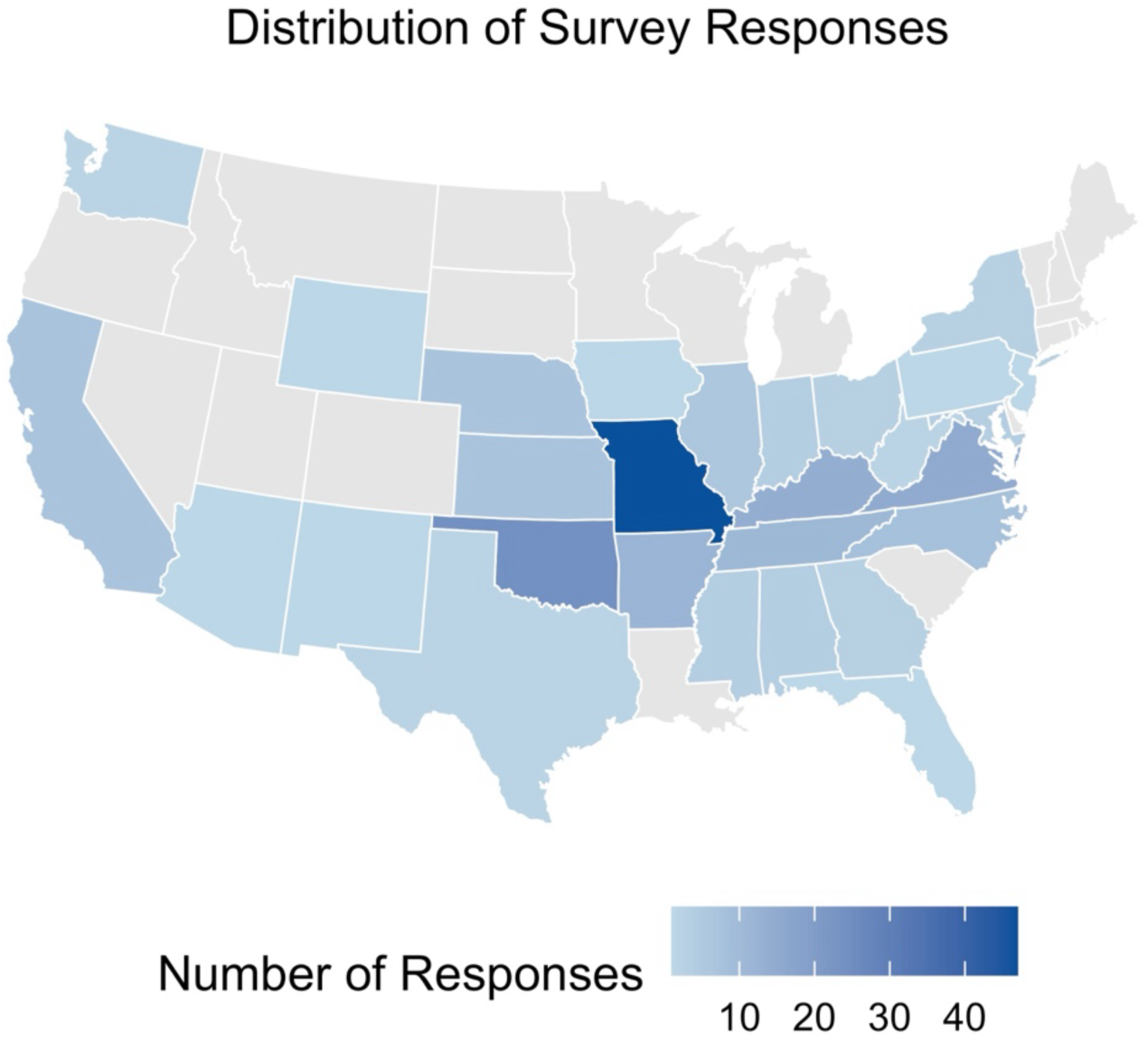
Distribution of survey responses from farmers and ranchers with self-identified alpha-gal syndrome. *Note: Responses were only received from the continental United States, therefore Alaska and Hawaii are not shown*.

**Table 1.**
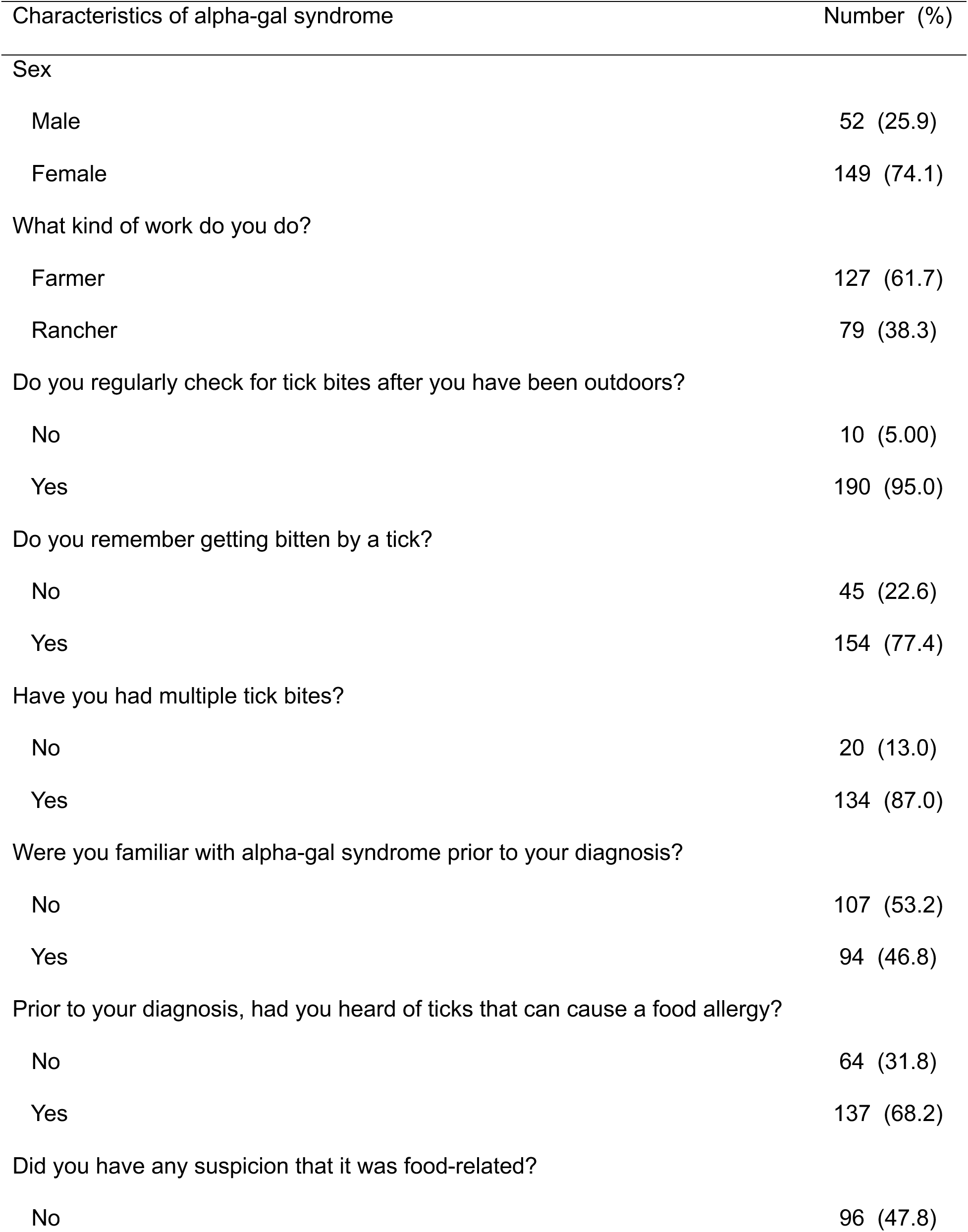
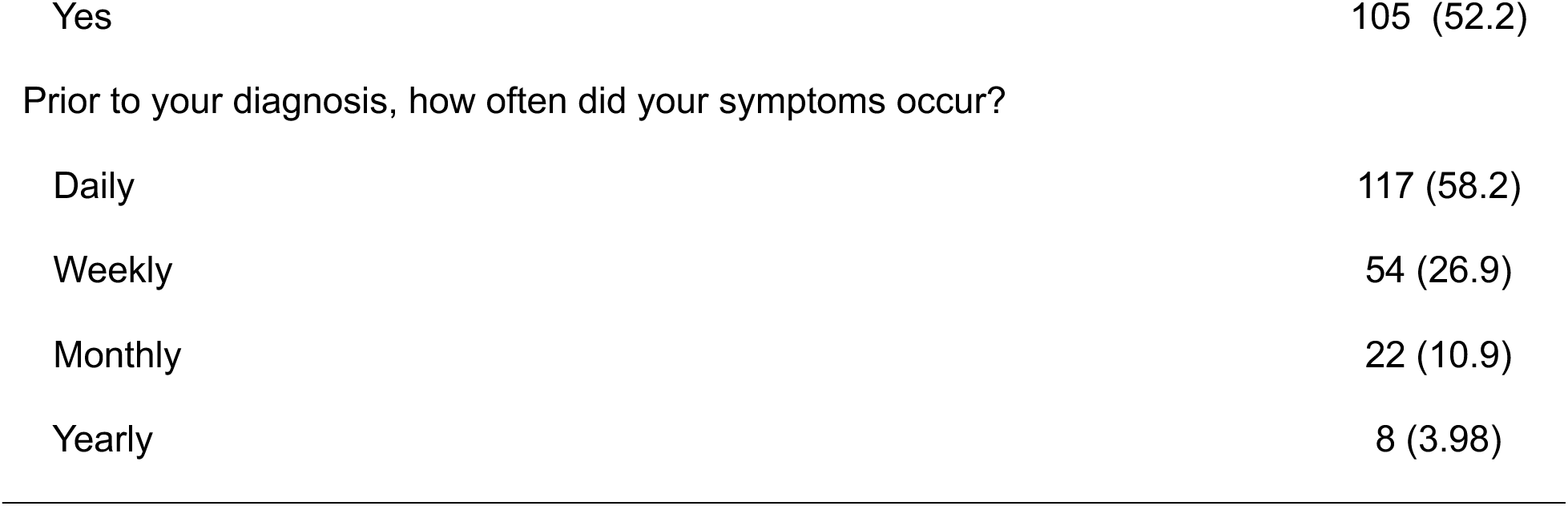
Characteristics of alpha-gal syndrome in 201 infected individuals, 2025.

| Characteristics of alpha-gal syndrome | Number (%) |
| --- | --- |
| Sex |  |
| Male | 52 (25.9) |
| Female | 149 (74.1) |
| What kind of work do you do? |  |
| Farmer | 127 (61.7) |
| Rancher | 79 (38.3) |
| Do you regularly check for tick bites after you have been outdoors? |  |
| No | 10 (5.00) |
| Yes | 190 (95.0) |
| Do you remember getting bitten by a tick? |  |
| No | 45 (22.6) |
| Yes | 154 (77.4) |
| Have you had multiple tick bites? |  |
| No | 20 (13.0) |
| Yes | 134 (87.0) |
| Were you familiar with alpha-gal syndrome prior to your diagnosis? |  |
| No | 107 (53.2) |
| Yes | 94 (46.8) |
| Prior to your diagnosis, had you heard of ticks that can cause a food allergy? |  |
| No | 64 (31.8) |
| Yes | 137 (68.2) |
| Did you have any suspicion that it was food-related? |  |
| No | 96 (47.8) |
| Yes | 105 (52.2) |
| Prior to your diagnosis, how often did your symptoms occur? |  |
| Daily | 117 (58.2) |
| Weekly | 54 (26.9) |
| Monthly | 22 (10.9) |
| Yearly | 8 (3.98) |

### Classification of reported symptoms

A multilevel latent class analysis (MCLA) was performed to understand how various reported symptoms grouped together (see *Methods*). The MLCA comparing model fit for up to six classes showed that at most four classes was the best solution, although the AIC and BIC did not always agree. The bootstrap likelihood ratio test (BLRT) with 1,000 bootstraps was used to provide evidence for the best model fit to the data. The two class model was superior to the one class model using AIC and BIC. The three class model showed a lower BIC than the four class model, but the entropy was below an acceptable level of 0.80. The results in **Table 2** argue for the four class model as the best fitting solution. Compared to the four class model, the five class model shows a slight decrease in the AIC, the BIC increased, the BLRT became much less statistically significant, and entropy was slightly lower. In the four class solution, Class one contained 36 individuals (17.9%), Class two 63 individuals (31.3%), Class three 50 individuals (24.9%) and Class four 52 individuals (25.9%).

**Table 2.**
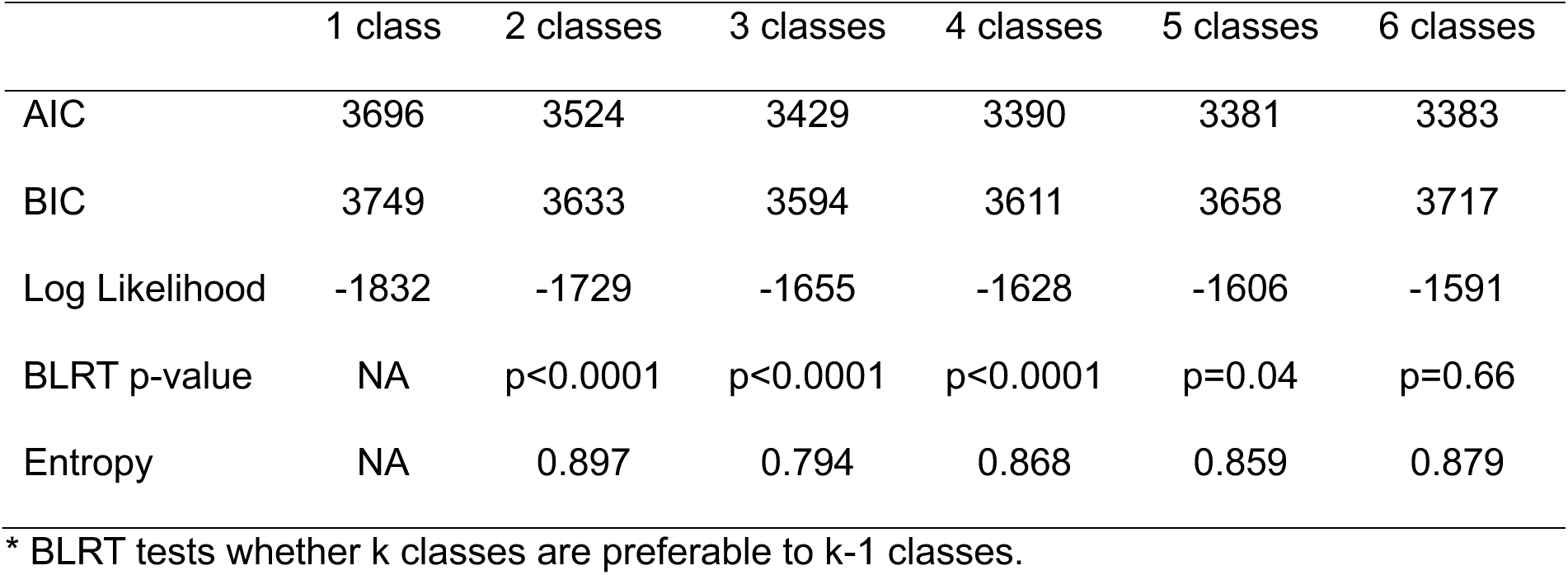
Model fit parameters for multilevel latent class analysis of 16 different symptoms reported by 201 with diagnosed alpha gal syndrome across the United States, 2025.

The classification probabilities for each of the four classes are shown below. The bolded diagonal elements represent classification agreement. Correct classification was observed at a probability of at least 91% (**Table 3**). Misclassification occurred less than 5% of the time in each of the other discordant categories.

**Table 3.**
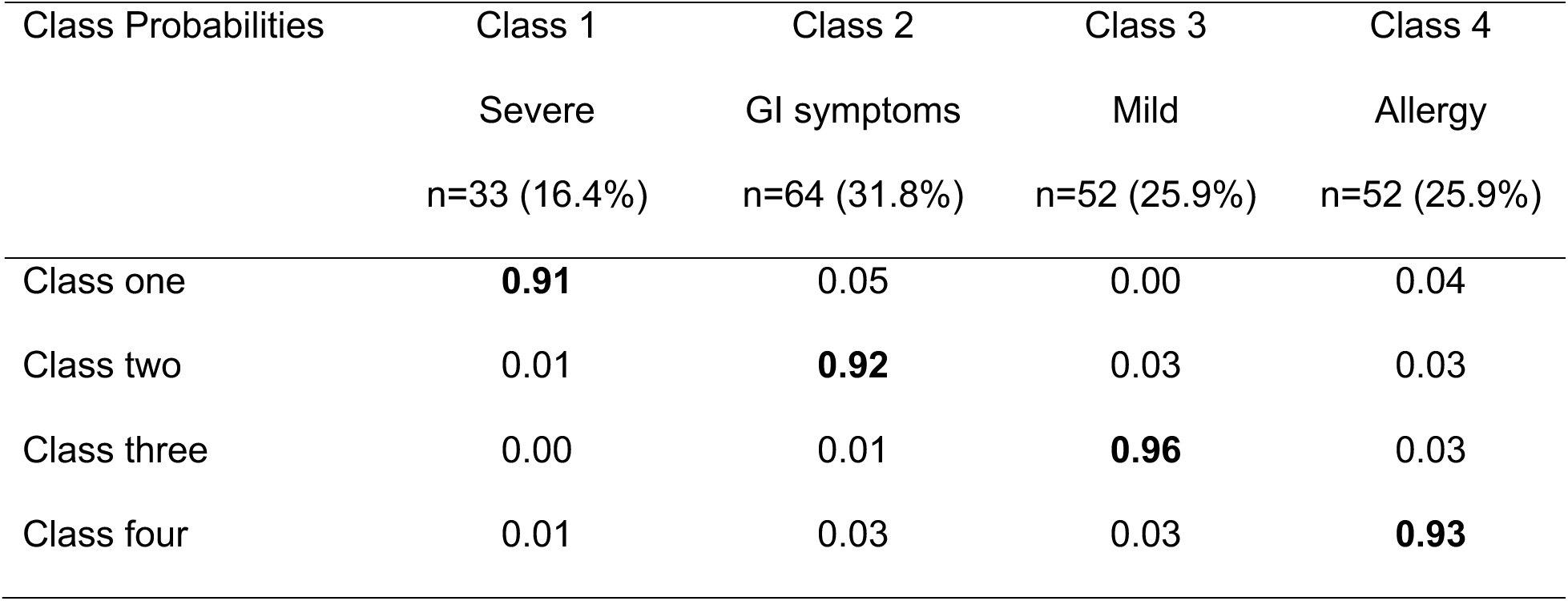
Classification accuracy in 201 individuals with alpha gal syndrome, 2025.

The four classes were not distinct in reporting of symptoms, as can be seen in the crossing of the lines in **Figure 2**. However, there were patterns in the symptoms. Class 1 was high on nearly all symptoms (labeled as “Severe”), although nausea and swelling were also frequently reported in Class 2 and Class 3. Class 2 primarily reported gastrointestinal symptoms such as abdominal cramps, diarrhea, nausea, and vomiting (labeled as “GI”). Class 3 was low on all symptoms but was highest on abdominal cramps, diarrhea, and difficulty breathing (labeled as “Mild”). Class 4 was dominated by allergic type reactions such as hives, itchy skin, and rash (labeled as “Allergy”). The symptoms reported in the “other” category that participants specifically reported such as fatigue, neurological symptoms (e.g. brain fog, seizures, headaches, dizziness), joint and muscle pain, and anaphylaxis did not show a high prevalence in any of the four classes; prevalences were less than 25% (**Table 4**). All symptoms specifically asked about were highly significantly different across classes (all p-values <0.0001). However, the symptoms in the “other” category were less so (fatigue p=0.04, neurological p=0.003, pain p=0.04) and anaphylaxis was not significantly different across the classes (p=0.17). The most severe group reported the fewest number of symptoms in the “other” category. The survey captured the primary symptoms that participants were experiencing, but a subgroup was experiencing fatigue, neurological effects, and muscle and joint pain. Anaphylactic reactions were relatively rare.

**Figure 2.**
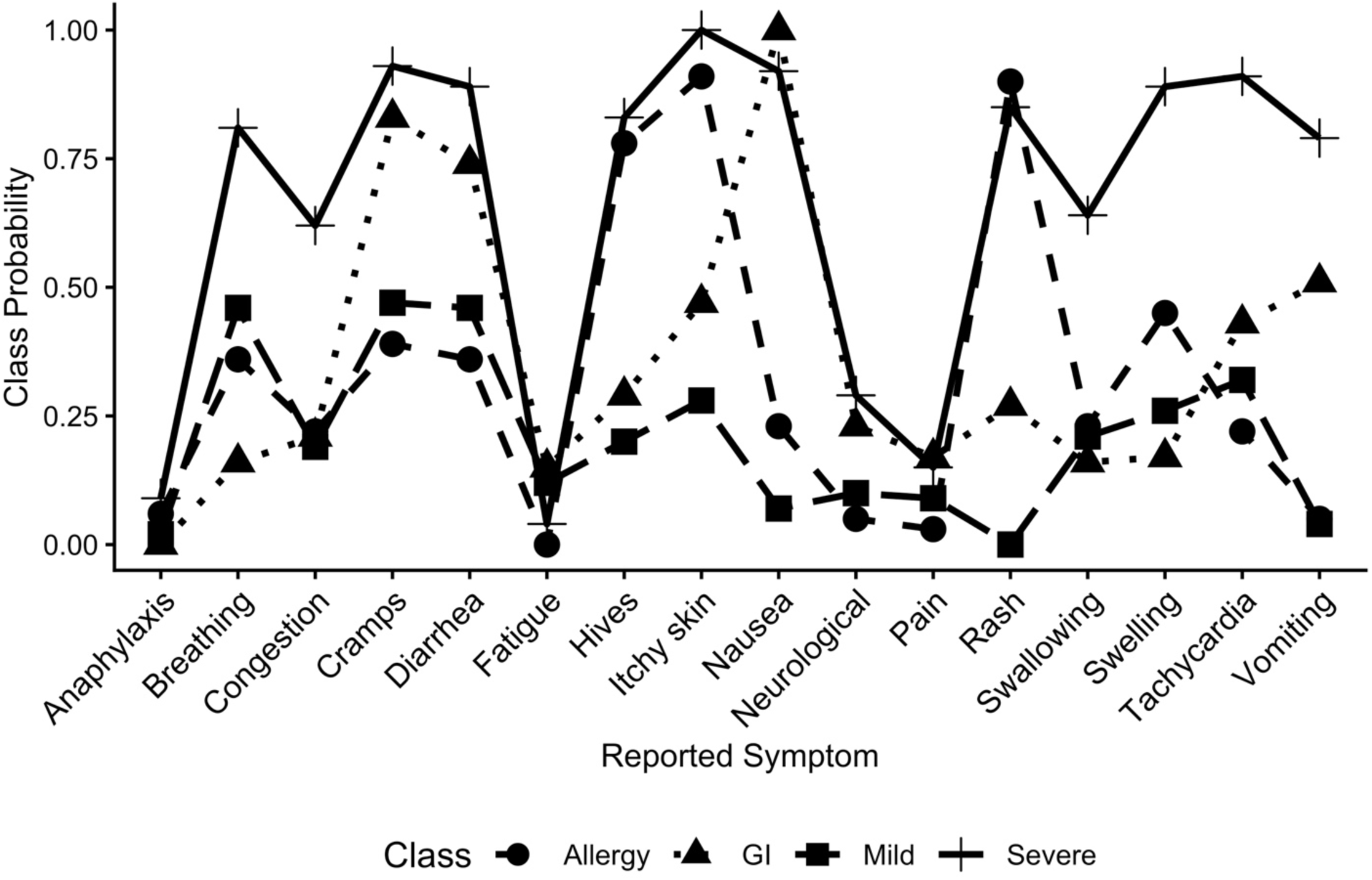
Probability of a symptom in each of the four classes in 201 participants with alpha gal syndrome, 2025.

**Table 4.** Prevalence of symptoms in total sample and for each class in 201 participants with alpha-gal syndrome, 2025.

| Symptom | Total Sample<br>(n=201) | Severe<br>(n=33) | GI<br>(n=64) | Mild<br>(n=52) | Allergy<br>(n=52) |
| --- | --- | --- | --- | --- | --- |
| Abdominal cramps | 128 (63.7) | 33 (91.7) | 53 (84.1) | 23 (46.0) | 19 (36.5) |
| Congestion | 56 (27.9) | 22 (61.1) | 12 (19.1) | 10 (20.0) | 12 (23.1) |
| Diarrhea | 119 (59.2) | 32 (88.9) | 47 (74.6) | 23 (46.0) | 19 (36.5) |
| Difficulty breathing | 81 (40.3) | 29 (80.6) | 10 (15.9) | 24 (48.0) | 18 (34.6) |
| Difficulty Swallowing | 55 (27.4) | 22 (66.1) | 11 (17.5) | 10 (20.0) | 12 (23.1) |
| Tachycardia | 86 (42.8) | 33 (91.7) | 28 (44.4) | 15 (30.0) | 10 (19.2) |
| Hives | 98 (48.8) | 30 (83.3) | 18 (28.6) | 9 (18.0) | 41 (78.9) |
| Itchy skin | 126 (62.7) | 36 (100) | 29 (46.0) | 13 (26.0) | 48 (92.3) |
| Nausea | 109 (54.2) | 33 (91.7) | 63 (100) | 2 (4.00) | 11 (21.1) |
| Rash | 94 (46.8) | 31 (86.1) | 16 (25.4) | 0 (0) | 47 (90.4) |
| Swelling | 78 (38.8) | 32 (88.9) | 10 (15.9) | 13 (26.0) | 23 (44.2) |
| Vomiting | 63 (31.3) | 29 (80.5) | 30 (47.6) | 2 (4.00) | 2 (3.85) |
| Fatigue | 17 (8.46) | 1 (2.78) | 10 (15.9) | 6 (12.0) | 0 (0) |
| Neurological | 32 (15.9) | 10 (27.8) | 15 (23.8) | 5 (10.0) | 2 (3.85) |
| Muscle and joint pain | 22 (10.9) | 5 (13.7) | 11 (17.5) | 5 (10.0) | 1 (1.92) |
| Anaphylaxis | 7 (3.48) | 3 (8.33) | 0 (0) | 1 (2.00) | 3 (5.77) |

### Participant characteristics in the four classes

In a chi-square test, class membership was significantly associated with sex (Χ^2^=19.6, p=0.0002). Females were more likely to be in the GI (abdominal cramps, diarrhea, and nausea) class (36.9%) compared to the severe class (20.8%), mild class (18.1%), or the allergy class (24.2%). Males were more likely to be members of the mild symptom class (44.2%) and less likely to be found in the severe class (9.62%), the GI class (15.4%), or the allergy class (30.8%).

Interestingly, those with prior knowledge of AGS were more often grouped into the GI class (37.2%) compared to the severe symptom group (7.95%), the mild symptom group (26.6%) or the allergy group (28.7%) (Χ^2^=13.5, p=0.004). Reporting of strong reactions to tick bites was associated with class membership (Χ^2^=14.7, p=0.002) where having a strong reaction to tick bites was highest in the GI class (23.9%), followed by the allergy class (19.4%), the mild symptom class (17.4%), and lowest in the severe class (7.46%).Type of agricultural operation, having had many tick bites, thinking the symptoms were food-related, or the frequency of symptoms was not associated with class membership.

**Table 5** shows the odds ratios and confidence interval from the generalized logistic regression of sex and familiarity with AGS on the four classes with allergy as the reference class. Females had a three-fold higher risk of being in the GI symptom class compared to males. Being familiar with AGS resulted in a 77% lower chance of being in the severe class compared to those unfamiliar. Reporting of symptoms may have been influenced by previous knowledge about AGS.

**Table 5.**
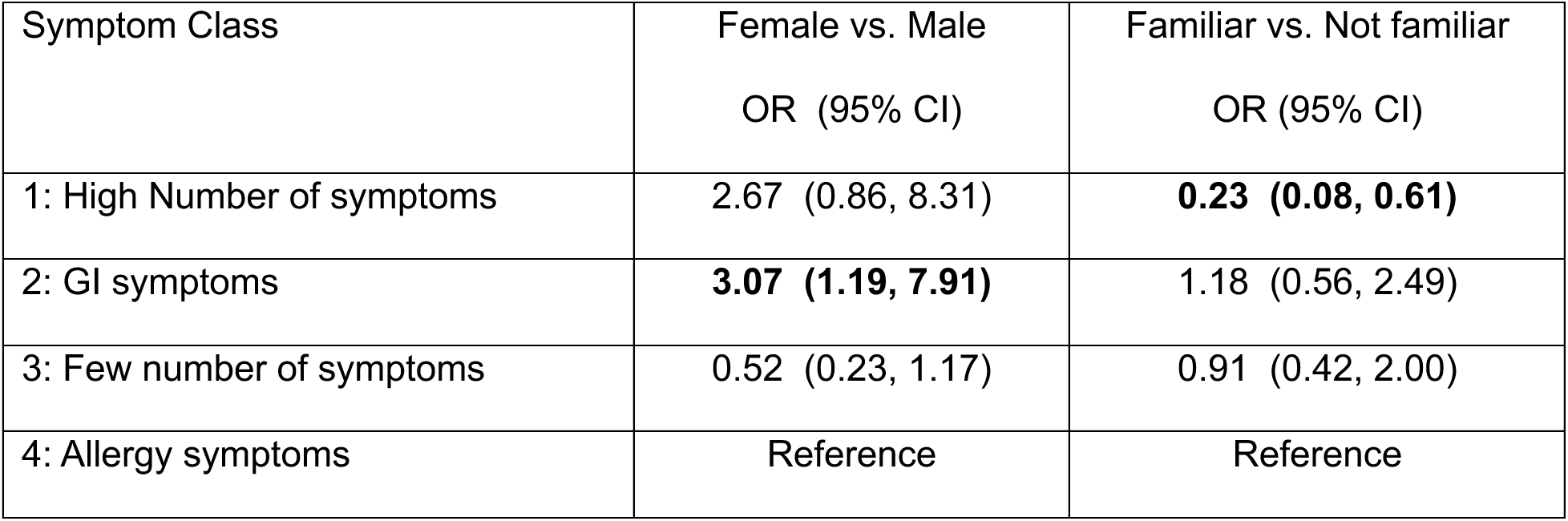
Associations between class membership and the explanatory variables sex and familiarity in 201 respondents diagnosed with AGS, 2025.

Age was not associated with class membership F[(3, 197)=1.13, p=0.34]. The mean age of members of the severe class was 47.5 (SD=11.3), the GI class 51.3 (SD=12.5), the mild symptom class 51.2 (SD=14.6), and the allergy class 47.8 (SD=15.5). Thinking the symptoms might be food-related, having a tick allergy, and having had multiple tick bites were not associated with any particular class. Having strong reactions to tick bites was not statistically significant with familiarity with AGS in the model, probably because 65% those with strong tick bite reactions were familiar with AGS.

### Diagnosis experience of those with AGS

The mean number of medical provider visits in 197 respondents was 2.98 (SD=2.23). For the 35 in the severe class, the mean number of visits was highest at 4.03 (SD=2.38), the 63 GI class members had 2.97 visits (SD=2.24), the 47 mild class members had 3.02 visits (SD=2.57), and the 52 in the allergy class had 2.25 visits (SD=1.41) to obtain a diagnosis. Compared to the allergy group, all regression coefficients were elevated, however, only the severe symptoms group reached statistical significance. The severe group had a 1.79 times higher number of expected medical provider visits than the allergy group (CI 1.32, 2.43). The GI group had a 1.32 times higher number of visits (CI 0.99, 1.76) and the mild group had a 1.34 times higher number of visits (CI 0.99, 1.82) than the allergy group. The group with only allergy symptoms required fewer medical provider visits to obtain a diagnosis.

Diagnosis occurred most often by a general practitioner (n=69, 34.3%), followed by an allergist (n=68, 33.8%), and a nursing professional (LPN, RN, NP) (n=18, 8.96). Other specialists that diagnosed AGS were gastroenterologists, dermatologists, ENT specialists, immunologists, neurologists, rheumatologist, and a few others medical professionals (hepatologist, infectious disease specialist, and physician assistants). Six cases were diagnosed by a nonmedical individual (2.99%) and four did not report who diagnosed them (1.99%).

## DISCUSSION

The primary goal of this study was to examine how AGS may be more pronounced among rural health communities, namely farmers and ranchers. To address this gap, we provide the first peer-reviewed study that captures the experiences of farmers and ranchers with AGS regarding their symptoms and initial diagnostic experience using a cohort of >200 participants.

Using a multilevel latent class analysis (MCLA) model (**Tables 1 and 2**) we examined how reported symptoms grouped together. We had a group who reported having nearly all of the symptoms asked about with high frequency and a second group that had these symptoms with low frequency. Two additional groups emerged where the dominant feature was gastrointestinal discomfort or allergic reaction, with little overlap between them. The classification into these four groups was accurate, with a greater than a 90% probability of being correct (**Table 3**). These identified groups require replication in additional studies and the risk factors related to these groups elucidated. Although few studies have assessed the commonalities and differences in reported symptoms, a 2018 study out of Cape Town with 84 AGS-positive individuals found that symptoms fell into two categories. Most (79%) had only abdominal cramping and 21% reported many symptoms including gastrointestinal, skin, and other serious symptoms.^25^ The range of symptoms asked about was relatively narrow but did include cardiovascular and respiratory symptoms, following strategies by other similar studies.^26^ Investigating how a constellation of diverse symptoms might fall into groups is more informative than asking about broad categories that include gastrointestinal and mucocutaneous disorders with abdominal cramping or hives since people have varying degrees of a wide range of symptoms, which complicates the ability to correctly diagnose AGS.

Symptomatology data was not collected on the study participants so it is unknown if the IgE levels correlated to symptom grouping. Prior studies however have identified IgE titers do not predict reaction severity.^27,28^ Genetic and immunological background (family or personal history of an autoimmune disorder) may be more deterministic in severity and grouping type than the level of IgE antibody. Nevertheless, further investigation into the relationship between IgE concentration and these symptom classes is needed.

A notable finding was sex-based differences in symptom classes. Women more often reported gastrointestinal discomfort and men were more likely to be in the mild symptom category (**Table 5**). Few studies have examined sex differences because they were conducted were in military personnel and in those who work primarily outdoors, male-dominated fields with few females. Sex differences were reported in a population-based, birth cohort study of 4,089 young adults in Stockholm Sweden.^29^ At the ages of 24, males were more likely to be IgE positive than females (8.9% vs. 3.4%, odds ratio = 2.8, 95% confidence interval 1.9-4.0). Males also reported a greater number of tick bites than females but there were no differences in IgE levels. The authors could not rule out the possibility that the males had greater exposure to the outdoors and to ticks than the females in the study. No other studies to our knowledge have reported differences in symptomatology by sex.

Of the participants, 46.8% were aware of AGS prior to their diagnosis (**Table 1**), a knowledge level relatively comparable to our survey among Indigenous bison workers^30^ and among healthcare providers.^22^ The participants who were familiar with alpha-gal were less likely to be in more severe category (**Table 5**). If they had familiarity with the symptoms of AGS, it might be expected that they would recognize and report greater symptoms. However, if they gained their knowledge of AGS because they had other tick-related conditions, they may not be able to separate out the various symptoms to their appropriate tick-borne cause and might report fewer AGS symptoms. This is information is best obtained in qualitative interviews which are now underway.

In a 2017 study of 28 individuals diagnosed with AGS, the mean number of medical encounters was 3.8, but the correct diagnosis was made only 9% of the time.^31^ We found that the average number of medical provider visits was 2.98 but were as high as ten or more visits (**Fig. 3A).** A correct diagnosis was eventually obtained in the participants here because our criteria for participation required self-reporting of having AGS. Therefore, this study does not tell us how many never received an AGS diagnosis despite having the condition. Also, similar to the Flaherty (2017) study,^31^ most of the diagnoses in the current study were made by a general practitioner or allergist (68%) but was often patient-driven. Unlike the Flaherty study, anaphylaxis was a rare event in the agricultural operators. Recruitment from a medical clinic may result in those with more serious symptoms compared to a population-based recruitment as performed here.

**Figure 3.**
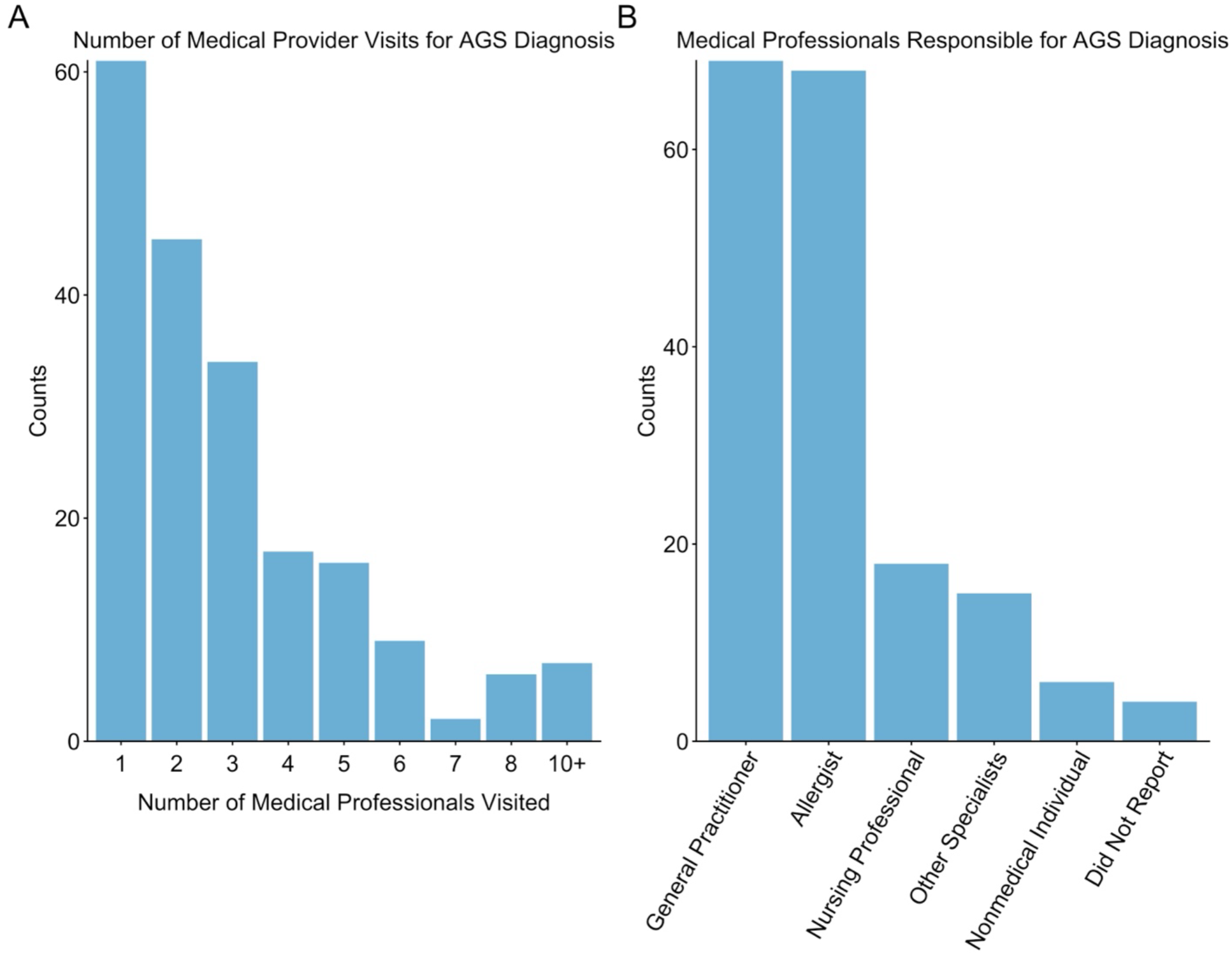
Farmer and rancher medical provider interactions during AGS diagnosis. A) Reported number of medical provider visits prior to receiving the AGS diagnosis. B) Frequency of medical professionals responsible for AGS diagnosis.

Three medical visits are costly and frustrating for the patient who is experiencing these symptoms. It is also a burden for farmers and ranchers in rural areas who have to drive far for medical treatment and lack access to medical specialists. As our results show, diagnosis is also a matter of the symptoms the medical provider is presented with. Diagnosis required fewer visits if the symptoms were allergy-related, which might lead to referral to an allergist who is more knowledgeable about AGS than a family/general medicine physician. Continued education and empowerment of both medical providers and patients is essential. A recent study reported that, nationally, 42% of health care providers had never heard of AGS and 35% who had heard of it said that they did not feel confident that they could diagnose it.^22^

Our study has limitations. Our population-based study has strengths because although it targeted farmers and ranchers which is male-dominated, most of the participants were females and females have been underrepresented in past studies of AGS. Our participants are limited to those who use primarily use social media and were aware of the Facebook Group and other platforms that shared the post. However, we promoted the survey in other ways, such as a network that is part of a NIOSH-funded agricultural health and safety center. We also asked about a wide range of symptoms and allowed participants to describe their symptoms in open-ended questions. In an effort to keep the survey short, we did not ask about a history of allergies or other chronic conditions, nor did we ask participants to provide any data confirming their AGS diagnosis. No previous studies have focused on these experiences in farmers and ranchers who are not only exposed more often to ticks due to the environment they work in,^15,17^ but may also be directly impacted by AGS from potential occupational tasks and exposures.^32^ This is a population where the impacts of AGS are both personal and economic.

This report contributes to the evolving AGS literature because even in a sample of 201, we provide evidence that the symptoms cluster in a pattern among those afflicted. We showed that diagnosis is easier when the symptom pattern looks like an allergy, which also aligns with studies showing the importance of atopy in some of these cases, but not all of them. Additionally, we provide evidence that knowledge is advancing in primary care and family physicians and they are getting better at either diagnosing AGS or knowing when to refer to an allergies to provide the appropriate testing. Collectively, such data can provide clear translation for appropriate outreach and education to empower at-risk individuals and medical providers for AGS.

### Funding Sources

This work was supported by funding provided by the Central States Center for Agricultural Safety and Health (CS-CASH) through a National Institute of Occupational Safety and Health (NIOSH) Agriculture, Forestry and Fishing Grant (U54OH010162).

### Disclosures

The authors have no disclosures or conflicts of interest to report.

## Data Availability

The data that support the findings of this study are available on request from the corresponding author. The data are not publicly available due to privacy or ethical restrictions.

## Acknowledgements

We would like to thank Sharon Forsyth (Alpha-gal Alliance) for her assistance with access and promotion to community groups for recruitment to this study. We sincerely thank all the farmer and rancher participants who provided their invaluable responses and assisted in this work.

## REFERENCES

1. Beard CB, Eisen L, Eisen RJ. The Rise of Ticks and Tickborne Diseases in the United States—Introduction. J Med Entomol. 2021;58(4):1487–1489. doi:10.1093/jme/tjab064

2. Gilbert L. The Impacts of Climate Change on Ticks and Tick-Borne Disease Risk. Annu Rev Entomol. 2021;66:373–388. doi:10.1146/annurev-ento-052720-094533

3. Sonenshine DE. Range Expansion of Tick Disease Vectors in North America: Implications for Spread of Tick-Borne Disease. Int J Environ Res Public Health. 2018;15(3):478. doi:10.3390/ijerph15030478

4. Thompson JM, Carpenter A, Kersh GJ, Wachs T, Commins SP, Salzer JS. Geographic Distribution of Suspected Alpha-gal Syndrome Cases – United States, January 2017-December 2022. MMWR Morb Mortal Wkly Rep. 2023;72(30):815–820. doi:10.15585/mmwr.mm7230a2

5. Commins SP, Platts-Mills TAE. Delayed Anaphylaxis to Red Meat in Patients with IgE Specific for Galactose alpha-1,3-Galactose (alpha-gal). Curr Allergy Asthma Rep. 2013;13(1):72–77. doi:10.1007/s11882-012-0315-y

6. Commins SP. Diagnosis & management of alpha-gal syndrome: lessons from 2,500 patients. Expert Review of Clinical Immunology. 2020;16(7):667–677. doi:10.1080/1744666X.2020.1782745

7. Crispell G, Commins SP, Archer-Hartman SA, et al. Discovery of Alpha-Gal-Containing Antigens in North American Tick Species Believed to Induce Red Meat Allergy. Front Immunol. 2019;10. doi:10.3389/fimmu.2019.01056

8. Sharma SR, Crispell G, Mohamed A, et al. Alpha-Gal Syndrome: Involvement of Amblyomma americanum α-D-Galactosidase and β-1,4 Galactosyltransferase Enzymes in α-Gal Metabolism. Front Cell Infect Microbiol. 2021;11:775371. doi:10.3389/fcimb.2021.775371

9. Butler WK, Oltean HN, Dykstra EA, Saunders E, Salzer JS, Commins SP. Onset of Alpha-Gal Syndrome after Tick Bite, Washington, USA. Emerg Infect Dis. 2025;31(4):829–832. doi:10.3201/eid3104.240577

10. Saunders EF, Sohail H, Myles DJ, et al. Alpha-Gal Syndrome after Ixodes scapularis Tick Bite and Statewide Surveillance, Maine, USA, 2014-2023. Emerg Infect Dis. 2025;31(4):809–813. doi:10.3201/eid3104.241265

11. Kennedy AC, Marshall E. Lone Star Ticks (Amblyomma americanum): Dela J Public Health. 2021;7(1):66–71. doi:10.32481/djph.2021.01.013

12. Eisen L. Tick species infesting humans in the United States. Ticks and Tick-borne Diseases. 2022;13(6):102025. doi:10.1016/j.ttbdis.2022.102025

13. Kersh GJ, Salzer J, Jones ES, et al. Tick bite as a risk factor for alpha-gal–specific immunoglobulin E antibodies and development of alpha-gal syndrome. *Annals of Allergy*, Asthma & Immunology. 2023;130(4):472–478. doi:10.1016/j.anai.2022.11.021

14. Wilson N, Vahey GM, McDonald E, et al. Tick bite risk factors and prevention measures in an area with emerging Powassan virus disease. Public Health Chall. 2023;2(4):e136. doi:10.1002/puh2.136

15. Roome A, Trombley D, Kern M, et al. Ticking Time Bomb: The Escalating Threat of Tick-Borne Diseases in Rural Farming Communities. J Agromedicine. 2026;31(1):102–108. doi:10.1080/1059924X.2025.2579639

16. Gual-Gonzalez L, Abiodun T, Nolan MS. Defining the tick-borne disease occupational risk among 4 high-risk vocations in South Carolina. J Med Entomol. 2025;62(3):712–717. doi:10.1093/jme/tjaf020

17. Piacentino JD, Schwartz BS. Occupational risk of Lyme disease: an epidemiological review. Occup Environ Med. 2002;59(2):75–84. doi:10.1136/oem.59.2.75

18. Schotthoefer A, Stinebaugh K, Martin M, Munoz-Zanzi C. Tickborne disease awareness and protective practices among U.S. Forest Service employees from the upper Midwest, USA. BMC Public Health. 2020;20(1):1575. doi:10.1186/s12889-020-09629-x

19. Kersh GJ, Salzer J, Jones ES, et al. Tick bite as a risk factor for alpha-gal–specific immunoglobulin E antibodies and development of alpha-gal syndrome. *Annals of Allergy*, Asthma & Immunology. 2023;130(4):472–478. doi:10.1016/j.anai.2022.11.021

20. Kaboli P, Blaine A, Mares J, Fortney J, Ono S, O’Shea AMJ. Health care access from the rural perspective: A narrative review. J Rural Health. 2026;42(1):e70119. doi:10.1111/jrh.70119

21. Javed A. Bridging the Health Care Gap in Rural Populations: Challenges, Innovations, and Solutions. The American Journal of Medicine. 2025;138(5):761–762. doi:10.1016/j.amjmed.2025.01.008

22. Carpenter A, Drexler NA, McCormick DW, et al. Health Care Provider Knowledge Regarding Alpha-gal Syndrome – United States, March-May 2022. MMWR Morb Mortal Wkly Rep. 2023;72(30):809–814. doi:10.15585/mmwr.mm7230a1

23. Celeux G, Soromenho G. An entropy criterion for assessing the number of clusters in a mixture model. Journal of Classification. 1996;13(2):195–212. doi:10.1007/BF01246098

24. Rochlin I, Egizi A, Ginsberg HS. Modeling of historical and current distributions of lone star tick, Amblyomma americanum (Acari: Ixodidae), is consistent with ancestral range recovery. Exp Appl Acarol. 2023;89(1):85–103. doi:10.1007/s10493-022-00765-0

25. Mabelane T, Basera W, Botha M, Thomas HF, Ramjith J, Levin ME. Predictive values of alpha-gal IgE levels and alpha-gal IgE: Total IgE ratio and oral food challenge-proven meat allergy in a population with a high prevalence of reported red meat allergy. Pediatr Allergy Immunol. 2018;29(8):841–849. doi:10.1111/pai.12969

26. Taylor ML, Kersh GJ, Salzer JS, et al. Intrinsic risk factors for alpha-gal syndrome in a case-control study, 2019 to 2020. Annals of Allergy, Asthma & Immunology. 2024;132(6):759–764.e2. doi:10.1016/j.anai.2024.01.029

27. Fischer J, Hebsaker J, Caponetto P, Platts-Mills TAE, Biedermann T. Galactose-alpha-1,3-galactose sensitization is a prerequisite for pork-kidney allergy and cofactor-related mammalian meat anaphylaxis. Journal of Allergy and Clinical Immunology. 2014;134(3):755–759.e1. doi:10.1016/j.jaci.2014.05.051

28. Commins SP, James HR, Stevens W, et al. Delayed clinical and ex vivo response to mammalian meat in patients with IgE to galactose-alpha-1,3-galactose. J Allergy Clin Immunol. 2014;134(1):108–115.e11. doi:10.1016/j.jaci.2014.01.024

29. Westman M, Asarnoj A, Ballardini N, et al. Alpha-gal sensitization among young adults is associated with male sex and polysensitization. J Allergy Clin Immunol Pract. 2022;10(1):333–335.e2. doi:10.1016/j.jaip.2021.10.018

30. Welch AM, Reuther M, Lynch-O’Brien LI, Samuelson MM, Cross ST. Prior knowledge and perceived impacts of tick-borne disease education among indigenous bison workers at an annual roundtable training. Front Public Health. 2026;13. doi:10.3389/fpubh.2025.1738923

31. Flaherty MG, Kaplan SJ, Jerath MR. Diagnosis of Life-Threatening Alpha-Gal Food Allergy Appears to Be Patient Driven. J Prim Care Community Health. 2017;8(4):345–348. doi:10.1177/2150131917705714

32. Nuñez-Orjales R, Martin-Lazaro J, Lopez-Freire S, Galan-Nieto A, Lombardero-Vega M, Carballada-Gonzalez F. Bovine Amniotic Fluid: A New and Occupational Source of Galactose-α-1,3-Galactose. J Investig Allergol Clin Immunol. 2017;27(5):313–314. doi:10.18176/jiaci.0170

